# Long-term outcomes of TAVI patients undergoing different pacing modality: LBBAP versus RVP

**DOI:** 10.1101/2024.06.12.24308735

**Authors:** Xi Wang, Yuanning Xu, Lijun Zeng, Kun Tan, Xueli Zhang, Xu Han, Tianyuan Xiong, Zhengang Zhao, Yong Peng, Jiafu Wei, Qiao Li, Sen He, Yong Chen, Minggang Zhou, Xi Li, Xin Wei, Yujia Liang, Wenxia Zhou, Lingyun Jiang, Xingbin Liu, Wei Meng, Rui Zhou, Guojun Xiong, Min Dai, Xiaojian Deng, Yuan Feng, Mao Chen

## Abstract

**Background:** New-onset permanent pacemaker implantation (PPMI) is still a common complication after transcatheter aortic valve implantation (TAVI) with adverse clinical outcomes. This study aims to investigate whether left bundle branch area pacing (LBBAP) improves long-term clinical results compared to traditional right ventricular pacing (RVP) in patients requiring PPMI following TAVI.

**Methods:** A total of 237 consecutive patients undergoing RVP (N=117) or LBBAP (N=120) following TAVI were retrospectively included. Long-term outcomes including all-cause death, heart failure rehospitalization (HFH) and left ventricular ejection fraction (LVEF) change compared to baseline were obtained until 5 years post-TAVI.

**Results:** The mean age of the overall population was 74 years with a mean surgical risk score as 4.4%. The paced QRS duration was significantly shorter in LBBAP group compared to RVP group (151 ± 18 vs. 122 ±12 ms, P<0.001). There was no difference between two groups in all-cause death (13.7% vs. 13.3%, adjusted HR: 0.76; 95% CI: 0.37 to 1.58; P=0.466) or the composite endpoint of death and HFH (29.9% vs. 19.2%, adjusted HR: 1.22; 95% CI: 0.70 to 2.13; P=0.476), however, the risk of HFH was significantly reduced in LBBAP group compared to RVP at 5 years after TAVI (21.4% vs. 7.5%, adjusted HR: 2.26; 95% CI: 1.01 to 5.08; P=0.048). There was a more marked evolution of LVEF over time in LBBAP group (P=0.046 for LVEF changes over time between groups).

**Conclusions:** LBBAP improved long-term clinical outcomes compared to RVP in patients undergoing PPMI after TAVI in terms of less HFH and better LVEF improvement.

## INTRODUCTION

Nowadays, transcatheter aortic valve implantation (TAVI) has become the standard- of-care for elderly patients with severe symptomatic aortic stenosis (AS) across all surgical risk categories[1,2]. Over the past decade, the risk of procedural-related complications has been largely reduced because of transcatheter heart valve (THV) design improvements as well as increased operator experience[3–5]. However, post- TAVI new-onset conduction disturbances and permanent pacemaker implantation (PPMI) are still common procedural complications[6]. Even though a patient-tailored pre-procedural planning together with optimized implantation techniques have been studied and conducted in most Heart Centers, the PPMI rate following TAVI remains a double-digit value, especially in TAVI using self-expanding THVs[7–9].

Previous studies have demonstrated the negative effect of post-TAVI PPMI on clinical outcomes, including increased risk of heart failure rehospitalizations (HFH) and lack of left ventricular function improvement, which is considered as a deleterious result of chronic right ventricular pacing (RVP)[6]. Recently, left bundle branch area pacing (LBBAP) has emerged as a novel physiologic pacing modality, showing excellent results for patients with conventional bradycardia pacing indications[10–12]. Meanwhile, feasibility and safety of LBBAP in TAVI patients has been demonstrated in several studies[13,14]. However, whether this novel pacing strategy could bring clinical benefits to patients undergoing TAVI remains under-investigated.

The aim of this study was to evaluate the outcome differences between RVP and LBBAP in a large cohort of patients requiring PPMI following TAVI, with long-term follow-up in both clinical endpoints and echocardiographic parameters.

## METHODS

### Study population

This was a multicenter, retrospective, observational study including all consecutive patients undergoing TAVI for severe AS with a permanent pacemaker implanted within 30 days after the procedure in the period April 2014 to May 2021 at 3 centers in China (West China Hospital, Mianyang Central Hospital, Deyang People’s Hospital). Exclusion criteria for this study were pre-procedural permanent pacemaker, valve-in-valve procedures, procedural failure (e.g., failure to valve implantation, conversion to open-heart surgery), procedural death and patients undergoing biventricular pacing (BVP). The enrolled patients were further divided according to the pacing modality (RVP vs. LBBAP groups). In accordance with local policies, all patients gave informed consent to the use of anonymous data for research. The Institutional Review Board of West China Hospital, Mianyang Central Hospital and Deyang People’s Hospital gave ethical approval for this work.

### PPMI after TAVI

Standardized TAVI procedure practice has been followed in each center. Patients received a PPMI following TAVI if there was a high-grade/complete atrioventricular block (AVB), severe symptomatic bradycardia, or if PPMI was deemed necessary by the electrophysiology team. LBBAP was introduced and widely adopted since April 2018 in participating centers. The technique for LBBAP has been previously described[15]. In brief, the Select Secure (model 3830, Medtronic Inc., Minneapolis, MN) pacing lead delivered through the C315HIS and C304His sheath (Medtronic Inc., Minneapolis, MN) was advanced and rotated into the muscular interventricular septum, finally positioned until the acceptable left bundle branch (LBB) area capture was achieved. LBBAP was considered successful if the unipolar paced QRS morphology demonstrated a Qr or qR pattern with any of the following: recording of LBB potential; demonstration of selective LBB/left ventricular (LV) septal capture; R- wave peak time in leads V5-6 <90 ms.

### Data collection & Follow-up

Baseline clinical characteristics, pre-procedural 12-lead electrocardiogram (ECG) and echocardiography results, TAVI procedural details and in-hospital outcomes were collected from the dedicated TAVI database in each center. Clinical follow-up was carried out at 30 days, 6 months, 12 months following TAVI and yearly afterwards. Since the adoption of LBBAP technique was much later than RVP in this study, all data and follow-up dates were censored after 5 years following TAVI, to eliminate the follow-up time difference between two groups. Long-term clinical outcomes include all-cause mortality, heart failure rehospitalizations and New York Heart Association (NYHA) classification, which were collected by several sources of information: outpatient clinical visits; phone contacts with patients and their families; and patient medical records provided from Health Care Big Data Center of Sichuan Province. For patients with multiple heart failure rehospitalizations, only the time point of the first episode was analyzed. TAVI procedural-related complications and all clinical events were defined according to the Valve Academic Research Consortium- 3 criteria[16].

Transthoracic echocardiography (TTE) results were available in all patients at baseline and discharge, and in 196 patients at ≥ 1-year follow-up. Left ventricular ejection fraction (LVEF) and left ventricular end-diastolic diameter (LVEDD) were calculated from the Simpson’s biplane method. Left ventricular dysfunction was defined as LVEF <50%. The paced QRS duration was measured at the ECG at the last follow up. Ventricular pacing threshold, sensing, impedance and pacing percentage were recorded from the last pacemaker interrogation at device clinic. Devices were programmed to achieve narrowest paced QRS duration while minimizing pacing burden.

### Statistical analysis

Continuous variables with normal distribution are expressed as mean ± SD and were compared using Student’s t-test; those without normal distribution are expressed as median (IQR) and were compared using the Wilcoxon rank sum test. Categorical data are presented as frequencies with percentages and were analyzed using the chi-square test, Fisher exact test, or Cochran-Armitage trend test. Within-group comparisons were performed by means of 2-tailed paired Student’s t-test (continuous data) or McNemar’s test (categorical data). Univariate and multivariable Cox proportional hazard models were used to estimate probability for the primary and secondary survival outcomes for the RVP and LBBAP groups. All multivariate models were adjusted for baseline differences in the univariate analysis including variables with a value of P < 0.10. The competing risk analysis for HFH with mortality as a competing risk was performed using the Aalen-Johansen method, and groups were compared using Gray’s test. A linear general model for repeated measures with interaction was used to compare the changes in LVEF at different time points between two groups. SPSS v.26 (IBM, USA) was used to perform all statistical analyses. Competing risk analysis was performed with the use of R version 4.3 (R Core Team, Austria). Analyses were considered significant at a two-tailed P value <0.05.

## RESULTS

### Study population

A total of 237 patients were included in this study, of which 117 patients underwent RVP following TAVI (RVP group) and 120 patients underwent LBBAP (LBBAP group). The overall study population had a mean age of 74 years and a mean Society of Thoracic Surgeons (STS) risk score as 4.4%. There were no significant differences in baseline features and comorbidities between two groups. The baseline LVEF was comparable (56 ± 13 vs. 57 ± 13 %, P=0.65), with 76 patients (32%) in the overall population presented with a baseline LVEF < 50%. Right bundle branch block was recognized on baseline ECG in 17 patients (7%), and the baseline QRS duration was similar in two groups (114 ± 21 vs. 111 ± 24 ms, P=0.52). An overview of all baseline clinical variables of both groups is shown in **Table 1**.

**Table 1.** Baseline characteristics.

|  | **RVP**<br>**N = 117** | **LBBAP**<br>**N = 120** | **P Value** |
| --- | --- | --- | --- |
| **Clinical characteristics** |  |  |  |
| Age, years | 74.7 ± 6.5 | 74.3 ± 6.6 | 0.64 |
| Male | 73 (62%) | 78 (65%) | 0.68 |
| Body Mass Index, kg/m<sup>2</sup> | 22.9 ± 3.4 | 23.3 ± 3.3 | 0.35 |
| Arterial hypertension | 59 (50%) | 67 (56%) | 0.40 |
| Diabetes mellitus | 24 (21%) | 29 (24%) | 0.50 |
| Coronary artery disease | 52 (44%) | 45 (38%) | 0.28 |
| Previous PCI | 14 (12%) | 23 (19%) | 0.13 |
| Prior cerebrovascular events | 16 (14%) | 20 (17%) | 0.52 |
| Chronic kidney disease | 12 (10%) | 9 (8%) | 0.46 |
| Chronic lung disease | 22 (19%) | 15 (13%) | 0.18 |
| NYHA class III-IV | 91 (78%) | 83 (69%) | 0.15 |
| STS risk score, % | 4.8 ± 3.0 | 4.1 ± 2.9 | 0.09 |
| **Echocardiography** |  |  |  |
| LVEF, % | 56 ± 13 | 57 ± 13 | 0.65 |
| LVEF < 50% | 43 (37%) | 33 (28%) | 0.13 |
| LVEDD, mm | 54.3 ± 7.2 | 55.0 ± 8.8 | 0.65 |
| Mean AV gradient, mmHg | 59.8 ± 19.1 | 48.3 ± 21.3 | <0.001 |
| Aortic regurgitation ≥ moderate | 45 (38%) | 58 (48%) | 0.14 |
| Mitral regurgitation ≥ moderate | 17 (15%) | 15 (13%) | 0.67 |
| **ECG** |  |  |  |
| Baseline QRS, ms | 114 ± 21 | 111 ± 24 | 0.52 |
| Atrial fibrillation | 20 (17%) | 23 (19%) | 0.68 |
| 1<sup>st</sup> AVB | 16 (14%) | 8 (7%) | 0.08 |
| LBBB | 5 (4%) | 3 (3%) | 0.69 |
| RBBB | 9 (8%) | 8 (7%) | 0.76 |

AV, aortic valve; AVB, atrioventricular block; ECG, electrocardiogram; LBBAP, left bundle branch area pacing; LBBB, left bundle branch block; LVEDD, left ventricular end-diastolic diameter; LVEF, left ventricular ejection fraction; NYHA, New York Heart Association; PCI, percutaneous coronary intervention; RBBB, right bundle branch block; RVP, right ventricular pacing; STS, Society of Thoracic Surgeons.

### TAVI procedural outcomes

All TAVI procedures were performed via transfemoral access. While the THV platform implanted during TAVI was different between two groups, with CoreValve (Medtronic, Minneapolis, MN) used in 15 patients only in RVP group, 169 patients (71%) of the overall population received a domestic VenusA – Valve (Venus Medtech, Hangzhou, China). TAVI procedural-related major complications were rare with similar rates between RVP and LBBAP group. At discharge, there was no significant difference in transvalvular mean gradient (12.5 ± 5.3 vs. 12.2 ± 6.1 mmHg, P=0.65) between two groups, and only 2 patients in each group presented with significant (≥ moderate) aortic regurgitation. Detailed TAVI procedural outcomes are shown in **Table 2**.

**Table 2.**
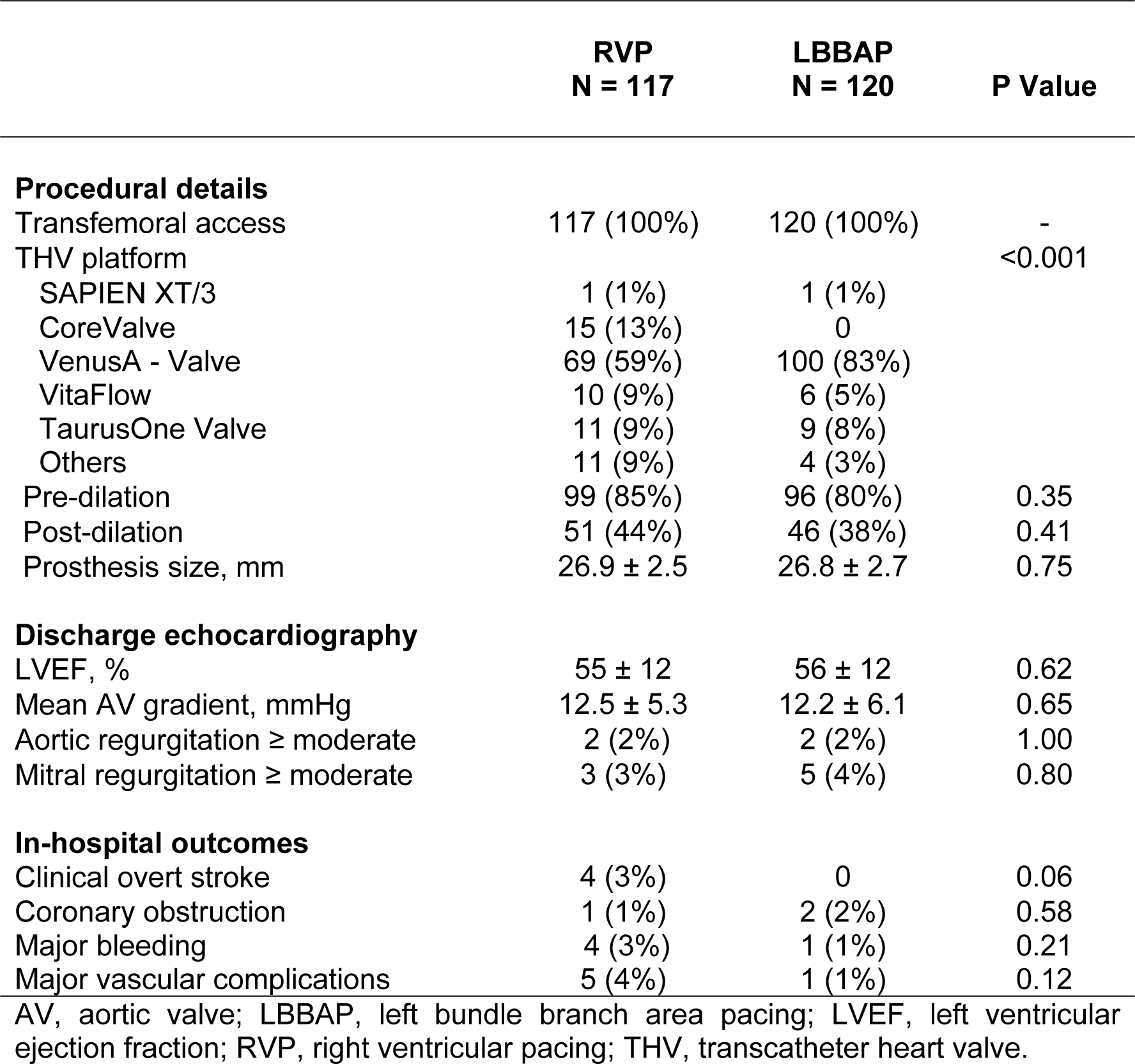
TAVI-procedural characteristics.

|  | **RVP**<br>**N = 117** | **LBBAP**<br>**N = 120** | **P Value** |
| --- | --- | --- | --- |
| **Procedural details** |  |  |  |
| Transfemoral access | 117 (100%) | 120 (100%) | - |
| THV platform |  |  | <0.001 |
| SAPIEN XT/3 | 1 (1%) | 1 (1%) |  |
| CoreValve | 15 (13%) | 0 |  |
| VenusA - Valve | 69 (59%) | 100 (83%) |  |
| VitaFlow | 10 (9%) | 6 (5%) |  |
| TaurusOne Valve | 11 (9%) | 9 (8%) |  |
| Others | 11 (9%) | 4 (3%) |  |
| Pre-dilation | 99 (85%) | 96 (80%) | 0.35 |
| Post-dilation | 51 (44%) | 46 (38%) | 0.41 |
| Prosthesis size, mm | 26.9 ± 2.5 | 26.8 ± 2.7 | 0.75 |
| **Discharge echocardiography** |  |  |  |
| LVEF, % | 55 ± 12 | 56 ± 12 | 0.62 |
| Mean AV gradient, mmHg | 12.5 ± 5.3 | 12.2 ± 6.1 | 0.65 |
| Aortic regurgitation ≥ moderate | 2 (2%) | 2 (2%) | 1.00 |
| Mitral regurgitation ≥ moderate | 3 (3%) | 5 (4%) | 0.80 |
| **In-hospital outcomes** |  |  |  |
| Clinical overt stroke | 4 (3%) | 0 | 0.06 |
| Coronary obstruction | 1 (1%) | 2 (2%) | 0.58 |
| Major bleeding | 4 (3%) | 1 (1%) | 0.21 |
| Major vascular complications | 5 (4%) | 1 (1%) | 0.12 |

AV, aortic valve; LBBAP, left bundle branch area pacing; LVEF, left ventricular ejection fraction; RVP, right ventricular pacing; THV, transcatheter heart valve.

### Pacing characteristics

Pacing characteristics at two groups are shown in **Table 3**. The most common indication for PPMI was high-degree or complete AVB (67%), and most (86%) pacemakers implanted were dual chamber. At the last follow up, the paced QRS duration was significantly shorter in LBBAP group (151 ± 18 vs. 122 ± 12 ms, P<0.001) **(Figure 1A)**. The ventricular pacing threshold was slightly higher in LBBAP group (0.68 ± 0.22 vs. 0.81 ± 0.32 V at 0.4 ms, P=0.02), while the sensing threshold (16.4 ± 4.6 vs. 17.8 ± 4.9 mV, P=0.21) and impedance (545.7 ± 99.0 vs. 567.9 ± 72.0 ohms, P=0.17) were comparable between two groups. The median of ventricular pacing percentage was over 95% in two groups (96.3 vs. 95.7%, P=0.45), while 70% of the overall population exhibited pacing > 40% of the time at last follow up.

**Figure 1.**
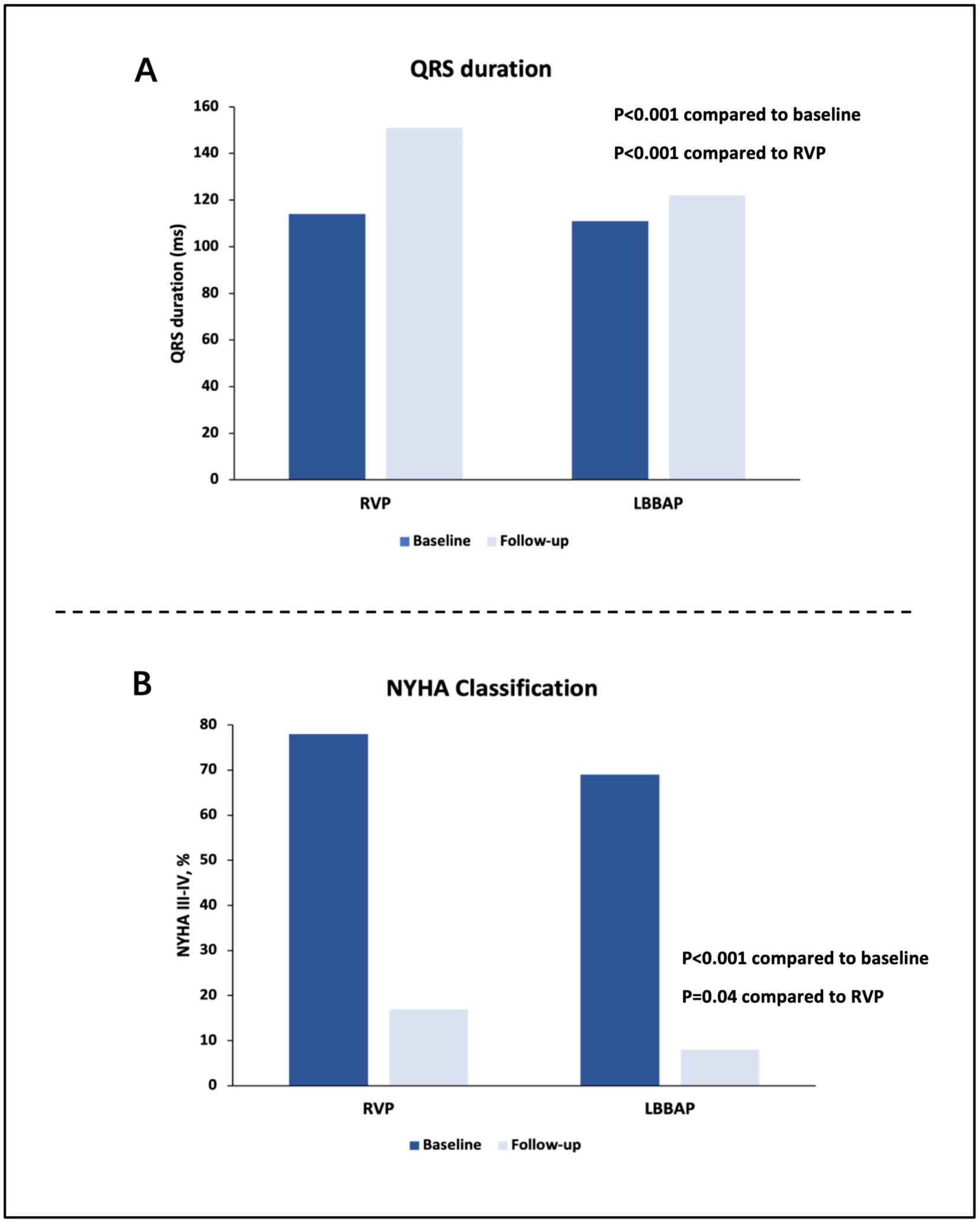
QRS duration and NYHA classification. Compared to patients in the RVP group, patients undergoing LBBAP presented with a significantly narrower paced QRS duration (A) (P<0.001) and more improved NYHA classification (B) (P=0.04) from baseline to the latest follow-up. LBBAP, left bundle branch area pacing; NYHA, New York Heart Association; RVP, right ventricular pacing.

**Table 3.** PPMI-procedural characteristics.

|  | **RVP**<br>**N = 117** | **LBBP**<br>**N = 120** | **P Value** |
| --- | --- | --- | --- |
| **Device type** |  |  |  |
| Single chamber | 17 (15%) | 16 (13%) | 0.79 |
| Dual chamber | 100 (85%) | 104 (87%) |  |
| **Indications** |  |  | 0.36 |
| Complete/high-degree AVB | 77 (66%) | 82 (68%) |  |
| LBBB+ 1<sup>st</sup> AVB | 27 (23%) | 25 (21%) |  |
| Severe symptomatic bradycardia | 11 (9%) | 7 (6%) |  |
| Alternating LBBB/RBBB | 2 (2%) | 6 (5%) |  |
| **Last follow-up** |  |  |  |
| Paced QRS duration, ms | 151 ± 18 | 122 ± 12 | <0.001 |
| Ventricular pacing threshold, V at 0.4 ms | 0.68 ± 0.22 | 0.81 ± 0.32 | 0.02 |
| Ventricular sensing threshold, mV | 16.4 ± 4.6 | 17.8 ± 4.9 | 0.21 |
| Ventricular impedance, ohms | 545.7 ± 99.0 | 567.9 ± 72.0 | 0.17 |
| Ventricular pacing, % | 96.3 (17.4,99.7) | 95.7 (29.2,99.9) | 0.45 |
| AVB, atrioventricular block; LBBAP, left bundle branch area pacing; LBBB, left bundle branch block; RBBB, right bundle branch block; RVP, right ventricular pacing. |  |  |  |

### Long-term clinical outcomes

At a median follow up of 48.5 (interquartile range: 34.9 – 60) months, a total of 32 patients died and 34 patients required HFH. Univariate and multivariable analysis, as well as the Kaplan-Meier curves for each clinical endpoint at 5-year (60-month) follow-up after adjusting for age, STS score and baseline LVEF are shown in **Figure 2** and **Central Illustration**. There was no difference in the rate of all-cause mortality between RVP and LBBAP group (13.7% vs. 13.3%, adjusted hazard ratio [HR]: 0.76; 95% confidence interval [CI]: 0.37 to 1.58; P=0.466). RVP was associated with a significantly higher risk of HFH (21.4% vs. 7.5%, adjusted HR: 2.26; 95% CI: 1.01 to 5.08; P=0.048). The competing risk analysis for HFH will all-cause mortality as a competing risk confirmed the significant decrease in HFH in LBBAP group compared to RVP (adjusted HR: 2.46, P=0.026). As for the composite endpoint of all-cause death and HFH, it occurred in more patients in RVP group, however, without reaching statistical significance on multivariable analysis (29.9% vs. 19.2%, adjusted HR: 1.22; 95% CI: 0.70 to 2.13; P=0.476). At the latest follow-up, NYHA classification improved significantly in both groups compared to baseline (portion of patients with NYHA III-IV: RVP: 78% vs.17%, P<0.001; LBBAP: 69% vs. 8%, P<0.001), while less patients in LBBAP group presented with NYHA class III-IV compared to RVP group (17% vs. 8%, P=0.04) (**Figure 1B**).

**Figure 2.**
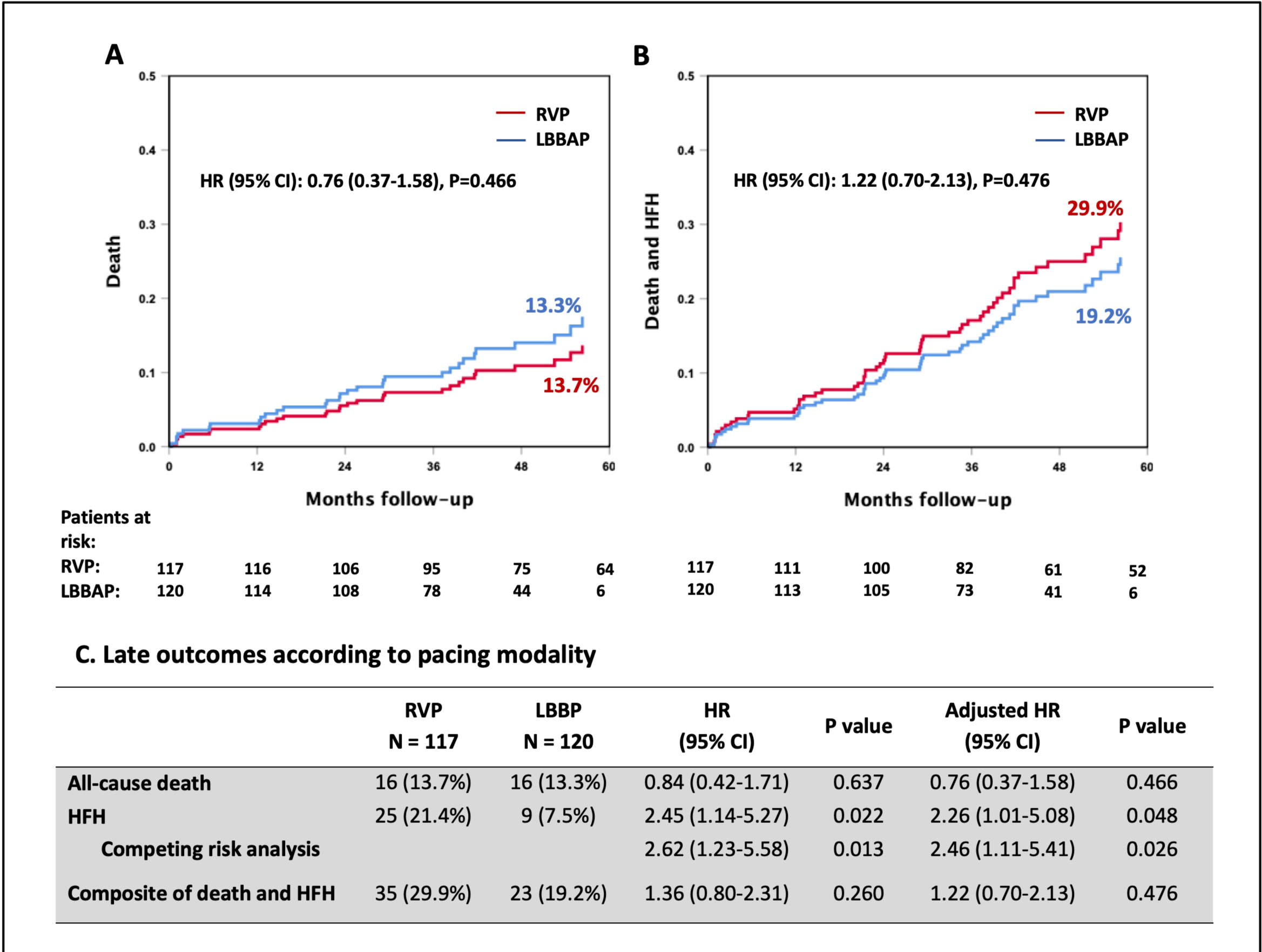
Long-term clinical outcomes according to pacing modality. Cumulative incidence curves at 5-year follow-up for (A) all-cause death and (B) composite endpoint of death or HFH. (C) Univariate and multivariable analysis of the clinical endpoints according to pacing modality (RVP vs. LBBAP), with the multivariable analysis adjusted for baseline variables. CI, confidence interval; HFH, heart failure rehospitalization; HR, hazard ratio; LBBAP, left bundle branch area pacing; RVP, right ventricular pacing.

### Echocardiographic change over time

TTE follow-up at least at one year after TAVI was available and analyzed in 196 patients (82.7%). At the latest follow-up, LVEDD decreased significantly in both groups compared to baseline (RVP: 54.3 ± 7.2 vs. 48.2 ± 6.4mm, P<0.001; LBBAP: 55.0 ± 8.8 vs. 48.9 ± 6.2mm, P<0.001), while LVEDD difference was similar between two groups (5.7 ± 7.6 vs. 5.6 ± 8.2mm, P=0.96). LVEF changes over time in all patients are shown in **Central Illustration.** LVEF increased over time in patients undergoing RVP and LBBAP following TAVI, with a mean LVEF of 59% and 63% at the latest follow-up, respectively. However, patients in LBBAP group experienced a more marked evolution of LVEF over time (P=0.046 for LVEF changes over time between groups). For patients with baseline left ventricular dysfunction, LVEF improved largely to a mean value of 53% and 57% in RVP and LBBAP group, respectively, while the LVEF recovery was not significantly different between the two groups (P=0.904 for LVEF changes over time between groups) (**Figure 3**).

**Figure 3.**
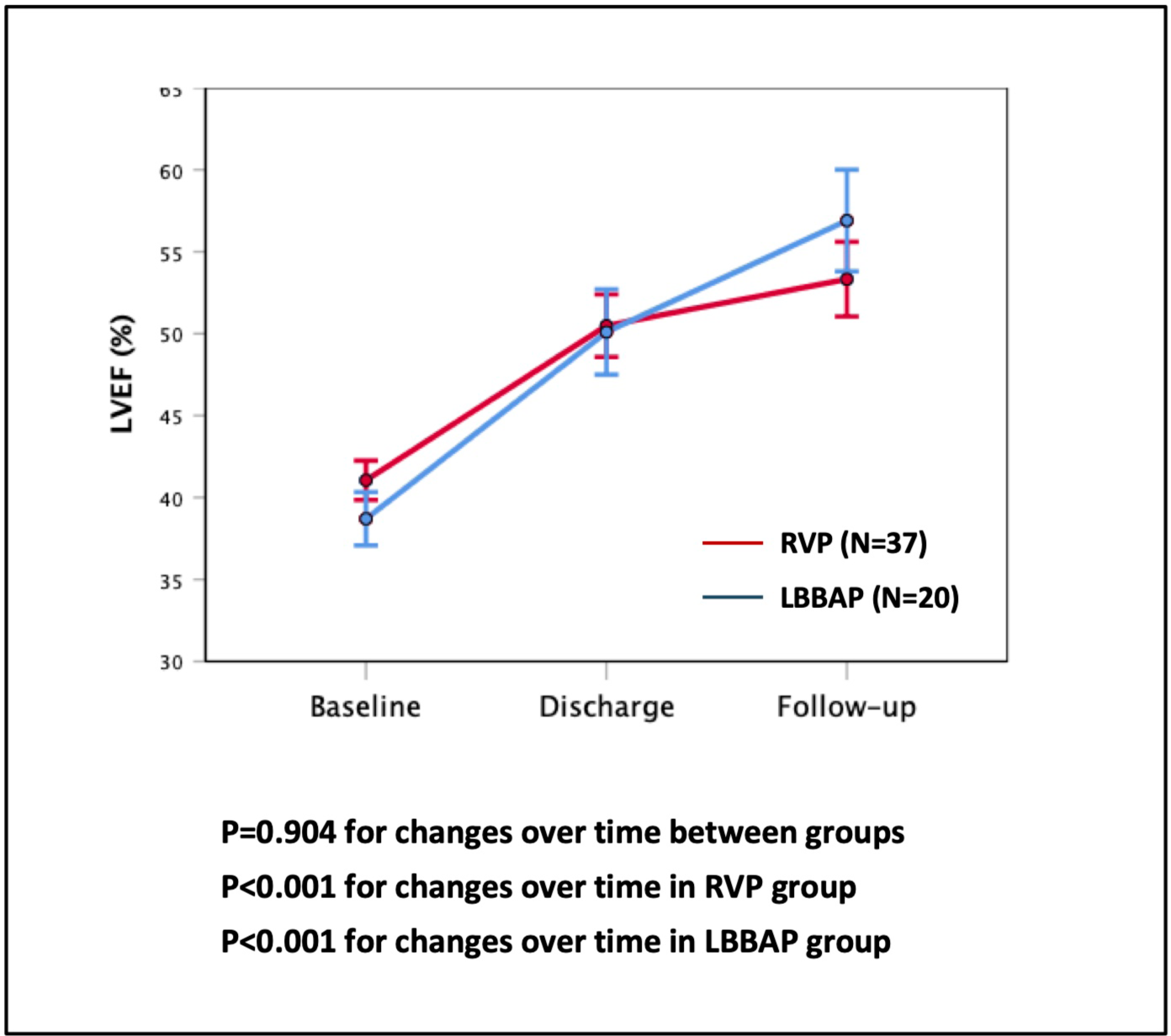
LVEF changes over time in patients with baseline left ventricular dysfunction. In patients with reduced left ventricular dysfunction at baseline (LVEF <50%), LVEF improved largely in both RVP and LBBAP group, while the LVEF recovery was not significantly different between the two groups (P=0.904 for LVEF changes over time between groups). LBBAP, left bundle branch area pacing; RVP, right ventricular pacing.

## DISCUSSION

To our knowledge, this is the first study reporting long-term clinical results in patients undergoing different pacing modalities following TAVI, with comparison between LBBAP and RVP in a large patient cohort. The main findings from this multicenter, retrospective observational study are as follows: (1) LBBAP was associated with a significant reduction in HFH in patients in need of PPMI after TAVI compared to RVP; (2) there was no significant difference in the incidence of all-cause death and the composite endpoint of all-cause death and HFH in the two groups; (3) there was a more marked LVEF improvement in LBBAP group compared to RVP group; (4) LBBAP resulted in narrower paced QRS duration than RVP.

It has been shown that PPMI has negative impacts on long-term clinical outcomes after TAVI, mostly due to the electrical and mechanical ventricular dyssynchrony caused by chronic RVP, which finally result in limited LVEF improvement and higher risk of HFH in these patients[17,18]. Coupled with increased patient costs and length of index hospitalization, the need of post-procedural PPMI is an obvious obstacle to further expanding the TAVI indication to younger patients with longer life expectancy. Previous studies mainly focus on risk factors of new-onset conduction disorders, as well as optimization of procedural strategy to lower the risk of post-procedural PPMI. However, PPMI are now still frequently needed after TAVI (>10% of patients using the new-generation self-expanding THVs), and the clinical prognosis in these patients urgently needs improvement[6].

Conduction system pacing, mainly utilizing His bundle pacing (HBP) and LBBAP, has been shown to achieve excellent electrical synchrony, and have the potential to prevent pacing-induced cardiomyopathy and the subsequent heart failure[19–21]. It is worth noticing that previous studies demonstrated a much higher feasibility of LBBAP than HBP in patients undergoing TAVI, characterized by higher success rate and lower pacing threshold[13,22]. This could be explained by the fact that self-expanding THV implantation results in extensive injury to the conduction system, which is hardly corrected via HBP. In our study, LBBAP was again proved to be feasible and effective in TAVI patients, with a success rate of 88% (120 in 137 patients who attempted LBBAP during the study period), accompanied by narrow paced QRS duration at long-term follow-up. The long-term pacing threshold of LBBAP was slightly higher than RVP (0.68 ± 0.22 vs. 0.81 ± 0.32 V at 0.4 ms, P=0.02), however, consistent with the threshold reported in previous literature[13].

Recently, several retrospective studies compared the clinical outcomes between LBBAP and RVP among patients undergoing PPMI. Sharma et al demonstrated that LBBAP resulted in improved outcomes compared to RVP in terms of the composite all-cause mortality, HFH, or upgrade to BVP (10.0% vs. 23.3%, HR 0.46; 95% CI 0.306–0.695; P <0.001) up to 3 years post procedure[23]. A meta-analysis from Leventopoulos et al also showed that LBBAP was associated with lower risk for HFH (RR:0.33, CI 95%:0.21 to 0.50; p < 0.001) and all-cause mortality (RR:0.52 CI 95%:0.34 to 0.80; p = 0.003) than RVP during a mean follow-up of 16 months[24]. However, few data have been published on the optimal pacing modality after TAVI, and long-term clinical outcomes using conduction system pacing following TAVI is missing. In the present study, a 5-year follow-up in a large cohort of patients undergoing LBBAP vs. RVP following TAVI was firstly provided, and LBBAP was associated with a lower risk of HFH, which is consistent with the clinical benefits of LBBAP reported in other patient cohorts. The incidence of the composite endpoint of all-cause death and HFH was not statistically different between LBBAP and RVP group, even though a lower incidence of this endpoint was shown in LBBAP, which was possibly affected by the similar all-cause mortality in both groups. It is worth mentioning that the majority of previous studies have failed to show any negative effect of PPMI on mortality in TAVI patients, therefore, LBBAP, as an optimized pacing modality, may not further benefit TAVI patients in terms of longer survival. More longer-term studies with a close follow-up of patients undergoing different pacing modalities after TAVI are warranted to provide solid evidence on the clinical benefits of LBBAP in TAVI patients.

The index TAVI procedure will result in LV reverse remodeling, shown as reduced LV volume/mass and improved LVEF after the procedure[25]. Previous studies have demonstrated a deleterious effect of PPMI on ventricular function among TAVI patients, mostly manifested as mildly decreased LVEF or limited LVEF improvement[6]. The current study showed a consistent finding in RVP group. Niu et al reported that LVEF in LBBAP group (N=20) was significantly higher than RVP group (N=30) (54.9 ± 6.7% vs. 48.9 ± 9.1%, P < 0.05) in TAVI patients at a mean follow-up of 15 months[26]. In this study, it’s also shown that LBBAP led to a more marked LVEF improvement over time from baseline to the latest follow-up (≥ 1 year), compared to RVP. The underlying mechanism of LVEF improvement was the maintenance of synchronous ventricular activation with LBBAP, which also contributed to the lower NYHA classification and reduced risk of HFH at long-term follow-up in LBBAP group. These findings have important clinical implications. A close collaboration between TAVI operators and electrophysiologists in the Heart Team is emphasized, with LBBAP considered as the first attempt instead of RVP in patients in need of PPMI after TAVI, especially those patients with potential high ventricular pacing burden (>20%), to optimize long-term outcomes of this patient cohort. And certainly, large multicenter randomized trials comparing RVP to LBBAP are expected in the future, to provide an impact on guideline recommendations of pacing modality in TAVI patients.

Patients who require PPMI after TAVI with baseline left ventricular dysfunction are a vulnerable population, with a possibly increased risk of pacing-induced heart failure and mortality[27]. Given the fact that LVEF still improves in these patients, cardiac resynchronization therapy (CRT) utilizing BVP is rarely performed at first PPMI in TAVI patients, and only considered when there is no LVEF recovery during follow-up. However, upgrade to BVP is associated with additional hospitalization, patient costs and risk of complications. A recent large observational study from Vijayaraman et al compared the clinical results between LBBAP and BVP in over 1700 patients undergoing CRT, and LBBAP was shown to be a reasonable alternative to BVP, with greater LVEF improvement and lower risk of HFH compared to BVP[28]. In this study, due to the small sample size, the LVEF recovery was not shown with difference between LBBAP and RVP group in patients with baseline left ventricular dysfunction. Future studies should focus on TAVI patients with reduced baseline LVEF, to determine whether LBBAP brings more benefits than RVP and upgrade to BVP in this specific population.

## LIMITATIONS

Important limitations of this study are its retrospective design, different time frame when applying RVP and LBBAP, and certain amount of loss of echocardiographic follow-up. Patients underwent LBBAP or RVP based on operator preference and were not randomized to either strategy. Due to the certain incidence of post-TAVI PPMI, it is hard to reach a particularly large cohort size within limited study time. However, the current study is the largest registry so far discussing pacing modality in TAVI population.

Another limitation of this study is that self-expanding THVs were predominantly implanted, and the THV types in both groups were different. This could possibly impact the clinical outcomes in both groups, which should be considered as a stratified variable in future randomized trials.

## CONCLUSIONS

LBBAP is a feasible and effective physiologic pacing modality for TAVI patients in need of permanent pacemaker. LBBAP was associated with significant reduction in HFH and more LVEF improvement compared to traditional RVP in TAVI patients at long-term follow-up (**Central Illustration**).

## ACKNOWLEDGEMENT

None.

## DATA AVAILABILITY STATEMENT

The data underlying this article will be shared on reasonable request to the corresponding author.

## ABBREVIATIONS

AS: aortic stenosis
AV: aortic valve
AVB: atrioventricular block
BVP: biventricular pacing
CI: confidence interval
CRT: cardiac resynchronization therapy
ECG: electrocardiogram
HBP: His bundle pacing
HFH: heart failure rehospitalization
HR: hazard ratio
LBB: left bundle branch
LBBAP: left bundle branch area pacing
LBBB: left bundle branch block
LV: left ventricular
LVEDD: left ventricular end-diastolic diameter
LVEF: left ventricular ejection fraction
NYHA: New York Heart Association
PCI: percutaneous coronary intervention
PPMI: permanent pacemaker implantation
RBBB: right bundle branch block
RR: relative risk
RVP: right ventricular pacing
STS: Society of Thoracic Surgeons
TAVI: transcatheter aortic valve implantation
THV: transcatheter heart valve
TTE: transthoracic echocardiography

---

**Central Illustration.**
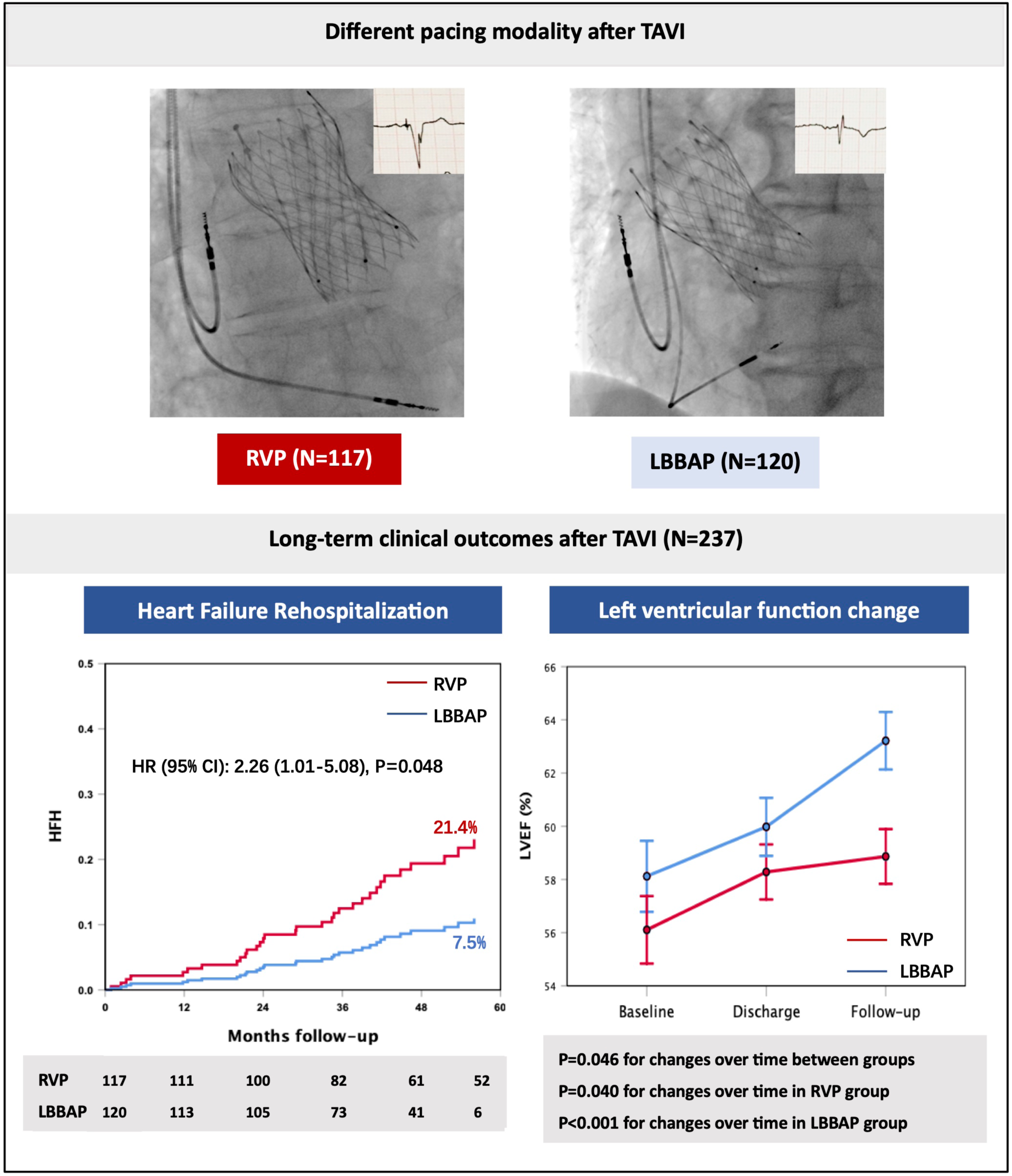
Long-term outcomes of LBBAP compared to RVP in TAVI patients. LBBAP is a feasible physiologic pacing modality for TAVI patients in need of permanent pacemaker, resulting in a narrow paced QRS duration. LBBAP was associated with significant reduction in HFH and more LVEF improvement compared to traditional RVP in TAVI patients at long-term follow-up. HFH, heart failure rehospitalization; LBBAP, left bundle branch area pacing; LVEF, left ventricular ejection function; RVP, right ventricular pacing; TAVI, transcatheter aortic valve implantation.

## REFERENCES

1. Writing Committee Members, Otto CM, Nishimura RA, et al. 2020 ACC/AHA Guideline for the Management of Patients With Valvular Heart Disease: Executive Summary: A Report of the American College of Cardiology/American Heart Association Joint Committee on Clinical Practice Guidelines. J Am Coll Cardiol. 2021;77:450–500.

2. Vahanian A, Beyersdorf F, Praz F, et al. 2021 ESC/EACTS Guidelines for the management of valvular heart disease. Eur Heart J. 2022;43:561–632.

3. Hellhammer K, Piayda K, Afzal S, et al. The Latest Evolution of the Medtronic CoreValve System in the Era of Transcatheter Aortic Valve Replacement: Matched Comparison of the Evolut PRO and Evolut R. JACC Cardiovasc Interv. 2018;11:2314–2322.

4. Meyer A, Unbehaun A, Hamandi M, et al. Comparison of 1-Year Survival and Frequency of Paravalvular Leakage Using the Sapien 3 Versus the Sapien XT for Transcatheter Aortic Valve Implantation for Aortic Stenosis. Am J Cardiol. 2017;120:2247–2255.

5. Jilaihawi H, Zhao Z, Du R, et al. Minimizing Permanent Pacemaker Following Repositionable Self-Expanding Transcatheter Aortic Valve Replacement. JACC Cardiovasc Interv. 2019;12:1796–1807.

6. Sammour Y, Krishnaswamy A, Kumar A, et al. Incidence, Predictors, and Implications of Permanent Pacemaker Requirement After Transcatheter Aortic Valve Replacement. JACC Cardiovasc Interv. 2021;14:115–134.

7. Aljabbary TF, Komatsu I, Ochiai T, et al. Cusp overlap method for self-expanding transcatheter aortic valve replacement. Catheter Cardiovasc Interv. 2024;103:202– 208.

8. Pascual I, Hernández-Vaquero D, Alperi A, et al. Permanent Pacemaker Reduction Using Cusp-Overlapping Projection in TAVR: A Propensity Score Analysis. JACC Cardiovasc Interv. 2022;15:150–161.

9. Wang X, Wong I, Bajoras V, et al. Impact of implantation technique on conduction disturbances for TAVR with the self-expanding portico/navitor valve. Catheter Cardiovasc Interv. 2022. Published onlineDecember 21, 2022. 10.1002/ccd.30517.

10. Zhang S, Zhou X, Gold MR. Left Bundle Branch Pacing: JACC Review Topic of the Week. J Am Coll Cardiol. 2019;74:3039–3049.

11. Cano Ó, Vijayaraman P. Left Bundle Branch Area Pacing: Implant Technique, Definitions, Outcomes, and Complications. Curr Cardiol Rep. 2021;23:155.

12. Su L, Wang S, Wu S, et al. Long-Term Safety and Feasibility of Left Bundle Branch Pacing in a Large Single-Center Study. Circ Arrhythm Electrophysiol. 2021;14:e009261.

13. Vijayaraman P, Cano Ó, Koruth JS, et al. His-Purkinje Conduction System Pacing Following Transcatheter Aortic Valve Replacement: Feasibility and Safety. JACC Clin Electrophysiol. 2020;6:649–657.

14. Wei H-Q, Li H, Liao H, et al. Feasibility and Safety of Permanent Left Bundle Branch Pacing in Patients With Conduction Disorders Following Prosthetic Cardiac Valves. Front Cardiovasc Med. 2021;8:705124.

15. Huang W, Chen X, Su L, Wu S, Xia X, Vijayaraman P. A beginner’s guide to permanent left bundle branch pacing. Heart Rhythm. 2019;16:1791–1796.

16. VARC-3 WRITING COMMITTEE:, Généreux P, Piazza N, et al. Valve Academic Research Consortium 3: Updated Endpoint Definitions for Aortic Valve Clinical Research. J Am Coll Cardiol. 2021;77:2717–2746.

17. Chamandi C, Barbanti M, Munoz-Garcia A, et al. Long-Term Outcomes in Patients With New Permanent Pacemaker Implantation Following Transcatheter Aortic Valve Replacement. JACC Cardiovasc Interv. 2018;11:301–310.

18. Costa G, Zappulla P, Barbanti M, et al. Pacemaker dependency after transcatheter aortic valve implantation: incidence, predictors and long-term outcomes. EuroIntervention. 2019;15:875–883.

19. Vijayaraman P, Chung MK, Dandamudi G, et al. His Bundle Pacing. J Am Coll Cardiol. 2018;72:927–947.

20. Wu S, Su L, Vijayaraman P, et al. Left Bundle Branch Pacing for Cardiac Resynchronization Therapy: Nonrandomized On-Treatment Comparison With His Bundle Pacing and Biventricular Pacing. Can J Cardiol. 2021;37:319–328.

21. Qu Q, Sun J-Y, Zhang Z-Y, et al. His-Purkinje conduction system pacing: A systematic review and network meta-analysis in bradycardia and conduction disorders. J Cardiovasc Electrophysiol. 2021;32:3245–3258.

22. Sharma PS, Subzposh FA, Ellenbogen KA, Vijayaraman P. Permanent His- bundle pacing in patients with prosthetic cardiac valves. Heart Rhythm. 2017;14:59– 64.

23. Sharma PS, Patel NR, Ravi V, et al. Clinical outcomes of left bundle branch area pacing compared to right ventricular pacing: Results from the Geisinger-Rush Conduction System Pacing Registry. Heart Rhythm. 2022;19:3–11.

24. Leventopoulos G, Travlos CK, Aronis KN, et al. Safety and efficacy of left bundle branch area pacing compared with right ventricular pacing in patients with bradyarrhythmia and conduction system disorders: Systematic review and meta- analysis. Int J Cardiol. 2023;390:131230.

25. Giannini C, Petronio AS, Nardi C, et al. Left ventricular reverse remodeling in percutaneous and surgical aortic bioprostheses: an echocardiographic study. J Am Soc Echocardiogr. 2011;24:28–36.

26. Niu H-X, Liu X, Gu M, et al. Conduction System Pacing for Post Transcatheter Aortic Valve Replacement Patients: Comparison With Right Ventricular Pacing. Front Cardiovasc Med. 2021;8:772548.

27. Maeno Y, Abramowitz Y, Israr S, et al. Prognostic Impact of Permanent Pacemaker Implantation in Patients With Low Left Ventricular Ejection Fraction Following Transcatheter Aortic Valve Replacement. J Invasive Cardiol. 2019;31:E15– E22.

28. Vijayaraman P, Sharma PS, Cano Ó, et al. Comparison of Left Bundle Branch Area Pacing and Biventricular Pacing in Candidates for Resynchronization Therapy. J Am Coll Cardiol. 2023;82:228–241.

